# Behavioral Telemetry for ICU Mortality Prediction: Documentation Pattern Analysis in 46,002 Low-Acuity MIMIC-IV Patients

**DOI:** 10.64898/2026.02.25.26347110

**Authors:** Greg Born

## Abstract

**Objective:** To develop and validate a predictive model incorporating behavioral telemetry signals—documentation pattern anomalies derived from routine EHR charting—alongside clinical variables for ICU mortality prediction in patients with low acute physiologic derangement.

**Materials and Methods:** Retrospective cohort study of 46,002 adult ICU stays from MIMIC-IV v3.1 (2008–2022) with SOFA scores 0–2, excluding neurological units. We extracted 66 variables spanning demographics, acuity, behavioral telemetry, clinical enrichment, and temporal factors. Progressive logistic regression models (M1–M7) were compared using cross-validation, DeLong tests, net reclassification improvement, and calibration analysis.

**Results:** Overall mortality was 9.34% (4,295 deaths). The clinical model (M5) achieved cross-validated AUROC 0.691 versus 0.639 for demographics alone (M2; ΔAUROC = 0.052, DeLong p = 4.41×10^−47^). NRI was 24.3%. Discordant care patients received 30.5% more chart events than concordant patients, with the sole deficit in neurological assessments (−15.4%), refuting the neglect hypothesis. Kaplan-Meier analysis confirmed survival separation (log-rank χ^2^ = 138.6, p = 5.32×10⁻^32^). In the most conservative subgroup (SOFA 0, no sedation, no ventilation, N = 11,158), orientation omission remained associated with mortality (adjusted OR 1.52, p = 0.027).

**Discussion:** Deep sedation and mechanical ventilation function as mediators on the causal pathway rather than traditional confounders; the discordant care signal retains significance after full sedation adjustment.

**Conclusion:** Documentation pattern analysis adds measurable predictive value for ICU mortality risk stratification and represents a novel signal for real-time EHR-based clinical decision support.

## Background and Significance

Early identification of patients at risk for clinical deterioration in the intensive care unit (ICU) remains a central challenge in critical care medicine. Traditional severity scoring systems— including SOFA,^2^ APACHE, and SAPS—rely on physiologic parameters and laboratory values, yet demonstrate limited discriminatory performance in patients with low measured acuity who nonetheless experience adverse outcomes. Among patients with SOFA scores of 0–2, representing those with minimal organ dysfunction by conventional criteria, mortality rates of 5– 10% suggest that current acuity measures fail to capture important risk signals.

We have previously demonstrated that documentation patterns in the electronic health record (EHR) contain latent signals of clinical significance. Specifically, the omission of orientation assessment—a cognitive engagement proxy—in otherwise thoroughly documented ICU patients predicts mortality with an adjusted odds ratio of 4.29 (E-value 8.04) at SOFA 0–2,^3^ while “discordant care”—the presence of extensive routine physical assessments without cognitive engagement assessment—carries an adjusted OR of 2.27 (E-value 3.97).^4^ Screening of 17 individual nursing assessments confirmed that orientation is the only assessment where omission predicts higher mortality.^5^ These behavioral telemetry signals are temporally stable across eras and amplified during night shifts (interaction OR 1.51, p = 3.04×10□□).^6^

The concept of behavioral telemetry extends beyond individual documentation omissions to a systems-level framework: what clinicians choose not to document may be as informative as what they do document. This negative telemetry principle has parallels in diagnostic error literature, where the absence of expected clinical actions frequently precedes adverse events,^7,16^ and in the missed nursing care literature, where care omissions predict patient outcomes^10^ and nurse staffing ratios are independently associated with mortality.^9^

The present study builds on these foundational observations with four objectives: (1) develop a predictive model integrating behavioral telemetry signals with clinical variables and newly available enrichment data (mechanical ventilation status, deep sedation, documentation velocity); (2) characterize the “more care but wrong care” phenomenon through documentation velocity analysis; (3) evaluate downstream outcomes through survival analysis and discharge disposition; and (4) address the sedation confounding question by isolating the behavioral signal in patients without sedation or mechanical ventilation.

## Methods

### Study Design and Data Source

This retrospective cohort study used data from MIMIC-IV v3.1, a publicly available critical care database comprising ICU stays at Beth Israel Deaconess Medical Center (2008–2022).1 Institutional review board approval was obtained by the database maintainers (MIT). Individual consent was waived due to the retrospective, de-identified nature of the data. This study reports a TRIPOD Type 1a prediction model development study.11 Reporting follows TRIPOD guidelines (Supplementary Figure S1).

### Study Population

We included adult ICU stays meeting: (1) age ≥18 years; (2) SOFA score 0–2 within the first 24 hours; (3) non-neurological ICU unit (excluding neuro-ICU and neuro-stepdown); and (4) ICU length of stay ≥24 hours. The resulting cohort comprised 46,002 ICU stays after merging with enrichment data. Some patients contributed multiple ICU stays; sensitivity analysis restricted to first stays showed consistent results.

### Outcome

The primary outcome was in-hospital mortality. Secondary outcomes included time-to-death (among decedents), discharge disposition (among survivors), and institutional discharge (composite of skilled nursing facility, long-term acute care, hospice, or rehabilitation facility).

### Predictor Variables

Variables were organized into progressive feature sets:

**Demographics and acuity (M2):** SOFA score (0–2), age (continuous), sex, Charlson comorbidity index.

**Behavioral telemetry (M3–M4):** Orientation assessment status (binary: documented vs. not documented within 24 hours), discordant care flag (routine care score ≥6 without orientation assessment), night shift indicator.

**Clinical enrichment (M5):** Deep sedation (any RASS ≤−3 in first 24 hours), mechanical ventilation status.

**Full behavioral (M6):** Addition of routine care score, individual assessment flags (RASS, GCS, turning, skin, Braden scale).

**Documentation velocity (M7):** Total chart events in 24 hours, unique items documented, category-specific event counts (vital signs, neurological, treatment, respiratory, pain/sedation, care plan). The complete variable dictionary with sources and model membership is provided in Supplementary Table S1.

### Statistical Analysis

Model development used 5-fold stratified cross-validation with logistic regression (sklearn LogisticRegression with StandardScaler preprocessing). Progressive models (M1–M7) were evaluated by cross-validated AUROC with standard deviations. Model comparison used DeLong tests for correlated AUROCs on full-sample predictions.^12^ Net reclassification improvement (NRI) used risk categories of <5%, 5–10%, 10–20%, and >20%.^13^ Calibration was assessed by Hosmer-Lemeshow goodness-of-fit and decile calibration plots using out-of-fold predictions.^14^

Machine learning models (random forest: 200 trees, max depth 6; gradient boosting: 200 estimators, max depth 4, learning rate 0.1) were compared on the clinical feature set (M5). Survival analysis used Kaplan-Meier estimation with log-rank tests, capped at 30 days. Cox proportional hazards regression was performed on the M5 feature set with Schoenfeld residual testing of the proportional hazards assumption; variables violating the assumption were addressed through stratification. Documentation velocity was compared using Welch’s t-tests. Discharge disposition used multivariable logistic regression adjusting for SOFA, age, sex, Charlson index, and night shift.

Sensitivity analyses included: (1) exclusion of patients with comfort care documentation; (2) restriction to SOFA 0; (3) restriction to the most conservative subgroup (SOFA 0, no deep sedation, no mechanical ventilation, N = 11,158) to isolate the behavioral signal from sedation confounding.

Analyses were performed in Python 3.12 with statsmodels, scikit-learn, and lifelines. Two-sided p < 0.05 defined statistical significance.

## Results

### Study Population

The cohort included 46,002 ICU stays with overall mortality of 9.34% (4,295 deaths). Mean age was 63.0 years, 55.2% male, mean SOFA 0.66, median Charlson comorbidity index 0 (mean 0.79, range 0–9). Mechanical ventilation was present in 40.9%, deep sedation (RASS ≤−3) in 50.6%, and CAM-ICU was assessed in only 2.4% of stays. Discordant care was identified in 19.3% (N = 8,891) with mortality of 14.4% versus 8.1% in concordant patients.

### Predictive Model Performance

Table 1 presents cross-validated AUROC for progressive models. SOFA alone (M1) achieved AUROC 0.534. Adding demographics (M2) increased performance to 0.639. Behavioral telemetry signals (orientation + discordant care, M3) added 0.015. The clinical model incorporating deep sedation and mechanical ventilation (M5) reached 0.691 (ΔAUROC vs M2 = 0.052). The enriched model with documentation velocity (M7) achieved 0.723 (ΔAUROC vs M2 = 0.083).

**Table 1.** Cross-Validated AUROC for Progressive Logistic Regression Models (5-Fold Stratified CV)

| Model | Features Added | N Vars | AUROC | SD | ΔAUROC vs M2 |
| --- | --- | --- | --- | --- | --- |
| M1: SOFA only | SOFA score | 1 | 0.534 | 0.011 | — |
| M2: Demographics | + Age, sex, Charlson | 4 | 0.639 | 0.005 | ref |
| M3: + Behavioral | + Orientation, discordant care | 6 | 0.654 | 0.006 | +0.015 |
| M4: + Night shift | + Night shift indicator | 7 | 0.662 | 0.007 | +0.022 |
| **M5: + Clinical** | + Deep sedation, mech. ventilation | 9 | **0.691** | 0.011 | **+0.052** |
| M6: + Full behavioral | + Routine care score, RASS, GCS, etc. | 15 | 0.695 | 0.010 | +0.055 |
| M7: + Doc velocity | + Chart events, category counts | 20 | 0.723 | 0.011 | +0.083 |

Abbreviations: AUROC, area under the receiver operating characteristic curve; SD, standard deviation; CV, cross-validation.

All model comparisons were statistically significant by DeLong test: M2 vs M5 (z = 14.4, p = 4.41×10□□□), M2 vs M7 (z = 20.9, p = 4.13×10□□□), M5 vs M7 (z = 16.2, p = 5.40×10□□□).

### Model Coefficients

Table 2 presents coefficients for the clinical model (M5). The strongest predictor was mechanical ventilation (OR 1.92, 95% CI 1.77–2.07), followed by discordant care (OR 1.59, 1.35–1.87), deep sedation (OR 1.50, 1.39–1.63), night shift (OR 1.47, 1.38–1.57), and orientation assessed (OR 1.47, 1.25–1.72). SOFA contributed modestly (OR 1.05 per point). Notably, the orientation coefficient reversed direction (became positive, indicating assessed patients had higher mortality) when deep sedation and mechanical ventilation entered the model—a confounding structure addressed in the Discussion. Full coefficients for all seven progressive models are presented in Supplementary Table S2.

**Table 2.**
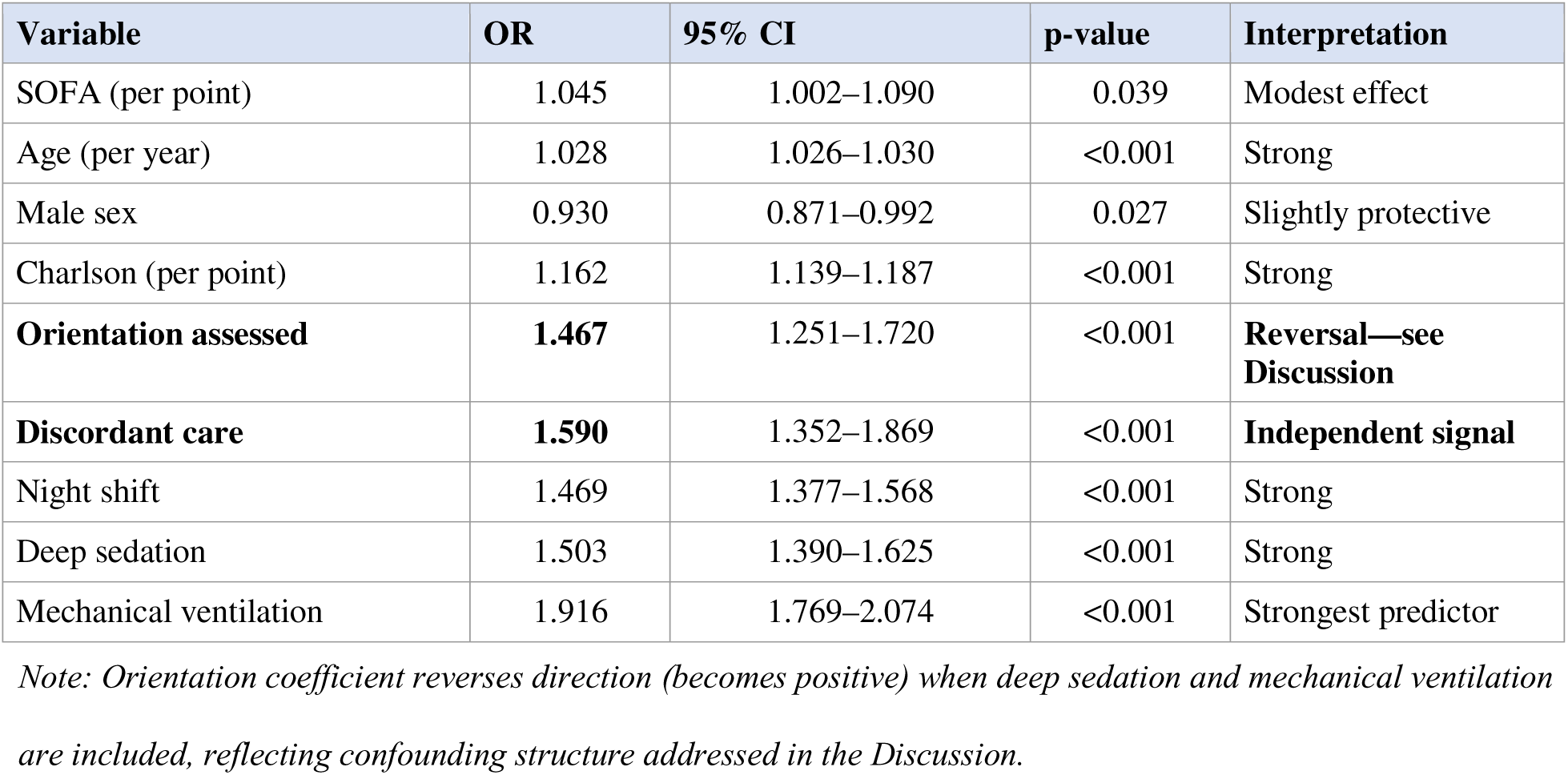
Logistic Regression Coefficients, Clinical Model (M5)

| Variable | OR | 95% CI | p-value | Interpretation |
| --- | --- | --- | --- | --- |
| SOFA (per point) | 1.045 | 1.002–1.090 | 0.039 | Modest effect |
| Age (per year) | 1.028 | 1.026–1.030 | <0.001 | Strong |
| Male sex | 0.930 | 0.871–0.992 | 0.027 | Slightly protective |
| Charlson (per point) | 1.162 | 1.139–1.187 | <0.001 | Strong |
| **Orientation assessed** | **1.467** | 1.251–1.720 | <0.001 | **Reversal—see Discussion** |
| **Discordant care** | **1.590** | 1.352–1.869 | <0.001 | **Independent signal** |
| Night shift | 1.469 | 1.377–1.568 | <0.001 | Strong |
| Deep sedation | 1.503 | 1.390–1.625 | <0.001 | Strong |
| Mechanical ventilation | 1.916 | 1.769–2.074 | <0.001 | Strongest predictor |

*Note: Orientation coefficient reverses direction (becomes positive) when deep sedation and mechanical ventilation are included, reflecting confounding structure addressed in the Discussion.*

### Reclassification and Calibration

The clinical model (M5) versus demographics (M2) yielded NRI of 24.3% (events 11.9%, non-events 12.4%) and IDI of 0.023. Calibration was adequate for both models (Hosmer-Lemeshow: M2 χ^2^ = 7.7, p = 0.46; M5 χ^2^ = 11.1, p = 0.20). The M5 model showed good spread across risk deciles, from 2.3% predicted risk (bottom decile) to 23.0% (top decile) with close agreement to observed rates (Figure 1D). Brier score improved from 0.0829 (M2) to 0.0810 (M5).

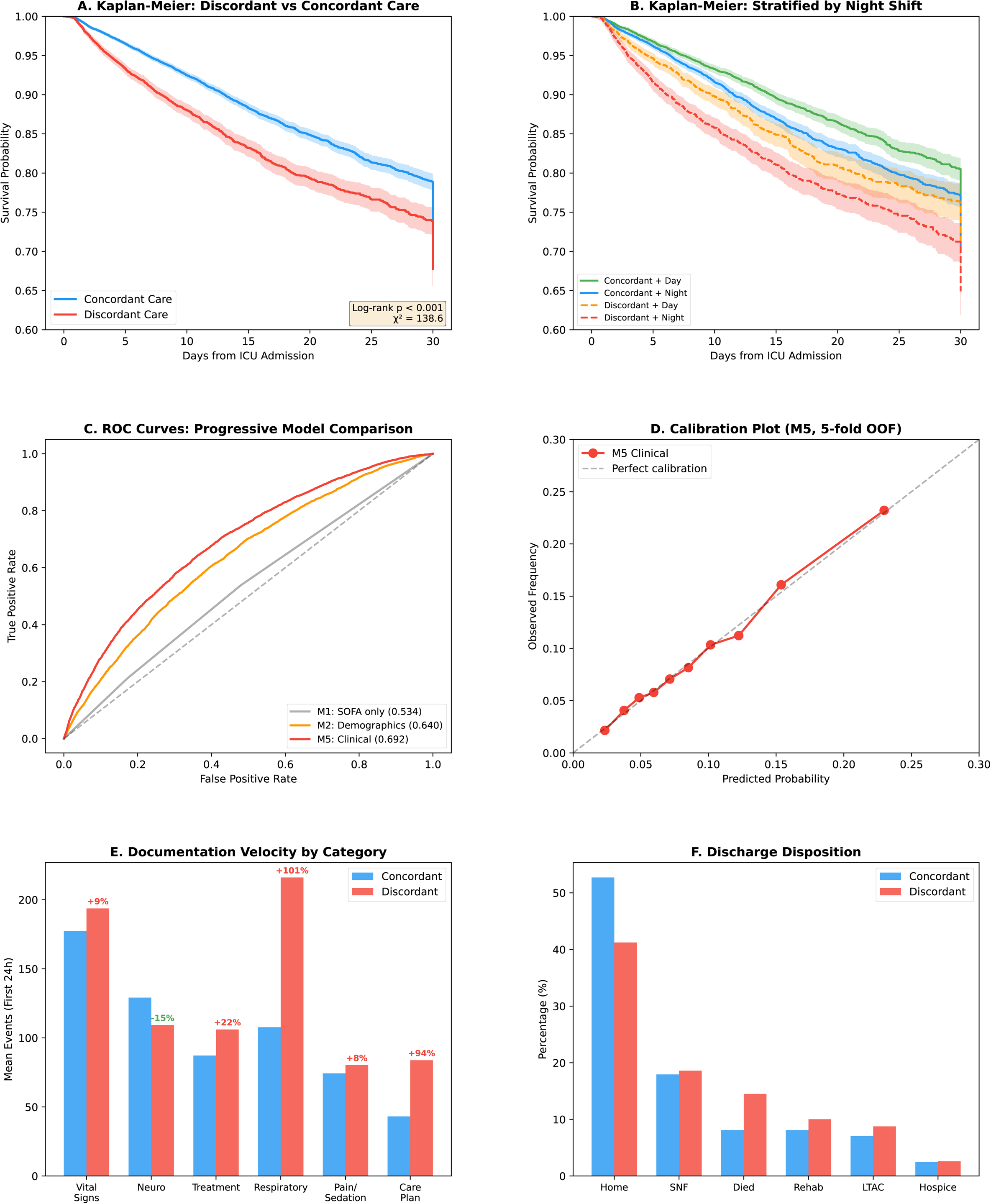

### Machine Learning Comparison

On the M5 feature set, random forest (AUROC 0.706) and gradient boosting (0.704) modestly outperformed logistic regression (0.691), suggesting mild non-linear relationships. Feature importance from gradient boosting identified respiratory events (0.309), age (0.163), documentation diversity (0.102), and neurological event counts (0.094) as the most discriminating features when documentation velocity was included (M7).

### Survival Analysis

Kaplan-Meier analysis demonstrated significant survival separation between discordant and concordant care groups (log-rank χ^2^ = 138.6, p = 5.32×10□^32^; Figure 1A). At 14 days, survival was 83.9% for discordant versus 89.2% for concordant (absolute difference 5.3%). Stratification by night shift revealed amplification: the discordant + night combination showed the steepest survival decline (χ^2^ = 86.2, p = 1.65×10□^2^□; Figure 1B). The compound risk group (SOFA 0 + night + discordant, N = 1,980) had 16.1% mortality versus 6.2% for the low-risk reference (SOFA 0 + day + concordant), a 2.6-fold difference (log-rank p = 2.80×10□¹□).

### Documentation Velocity: The “More Care, Wrong Care” Phenomenon

Discordant care patients received significantly MORE documentation than concordant patients across nearly all assessment categories (Table 3). Total chart events in the first 24 hours were 1,558 for discordant versus 1,194 for concordant (+30.5%, p < 0.001). Respiratory events showed the largest excess (+100.7%), followed by care plan events (+94.3%) and treatment events (+21.6%).

**Table 3.** Documentation Velocity by Category (First 24 Hours)

| Category | Discordant | Concordant | Difference | % Diff | p-value |
| --- | --- | --- | --- | --- | --- |
| **Total events** | 1,558 | 1,194 | +365 | **+30.5%** | <0.001 |
| Vital signs | 194 | 177 | +16 | +9.2% | <0.001 |
| **Neuro (SOLE DEFICIT)** | **109** | 129 | **-20** | **-15.4%** | <0.001 |
| Treatment | 106 | 87 | +19 | +21.6% | <0.001 |
| Respiratory | 216 | 108 | +108 | +100.7% | <0.001 |
| Pain/sedation | 80 | 74 | +6 | +8.1% | <0.001 |
| Care plan | 84 | 43 | +41 | +94.3% | <0.001 |

*Values are mean events per stay. Neurological events (bold) represent the sole category where discordant patients have fewer events. All comparisons $p < 0.001$ by Welch’s *t*-test.*

**Table 4.** Clinical Characteristics of Discordant vs. Concordant Patients.

| Characteristic | Discordant (N=8,891) | Concordant (N=37,111) | p-value |
| --- | --- | --- | --- |
| In-hospital mortality | 14.4% | 8.1% | <0.001 |
| Deep sedation (RASS $\leq -3$) | **90.5%** | 41.0% | <0.001 |
| Mechanically ventilated | **88.2%** | 29.5% | <0.001 |
| CAM-ICU assessed | 0.1% | 2.9% | <0.001 |
| RASS assessments (mean) | 6.0 | 4.7 | <0.001 |
| Institutional discharge (survivors) | 46.7% | 38.7% | <0.001 |
| Median hospital LOS (days) | 8.7 | 7.3 | <0.001 |

RASS, Richmond Agitation-Sedation Scale; CAM-ICU, Confusion Assessment Method for the ICU; LOS, length of stay.

The sole exception was neurological events, where discordant patients received 15.4% FEWER events (109 vs 129, p < 0.001; Figure 1E). This “more care but wrong care” pattern persisted among decedents: discordant patients who died received 1,633 chart events versus 1,257 for concordant decedents (+375 events, +29.9%).

### Time-to-First-Orientation

Among patients who received orientation assessment, the median time to first documentation was 1.1 hours (mean 3.1 hours). A U-shaped mortality pattern emerged by timing bin: patients assessed at 0–2 hours had 8.4% mortality, those assessed at 4–6 hours had 6.7%, those at 8–12 hours had 3.7% (the lowest), and those without any traceable orientation charting event (N = 4,737) had 24.7% mortality. In regression, log-transformed time to first orientation yielded OR 0.754 (95% CI 0.717–0.793, p = 3.03×10□^2^□). The U-shape is consistent with early assessment capturing acutely ill patients, late assessment reflecting survivorship selection, and never- assessed reflecting care process failure.

### Discharge Disposition

Among survivors (N = 41,707), discordant care was associated with lower home discharge (48.1% vs 57.3%) and higher institutional discharge (46.7% vs 38.7%; Figure 1F). The adjusted OR for institutional discharge was 1.50 (95% CI 1.42–1.58, p = 3.20×10□□□), indicating that behavioral telemetry signals predict not only mortality but also functional outcomes. Hospital length of stay was longer for discordant patients (median 8.9 vs 7.2 days, Mann-Whitney p = 1.21×10 ¹^33^).

### Time-to-Event Analysis

Cox proportional hazards regression on the M5 feature set confirmed the cross-sectional findings in a time-to-event framework. Discordant care was associated with a hazard ratio (HR) of 1.49 (95% CI 1.28–1.73, p < 0.005), orientation assessed HR 1.39 (1.19–1.61, p < 0.005), and mechanical ventilation HR 1.52 (1.41–1.63, p < 0.005). Model concordance was 0.67. Proportional hazards testing confirmed that the key behavioral variables (orientation assessment, discordant care) satisfied the PH assumption (p = 0.46 and p = 0.08, respectively), while deep sedation, mechanical ventilation, and night shift showed violations (p < 0.001, p < 0.001, and p = 0.007, respectively). Stratified Cox regression (stratifying on deep sedation and mechanical ventilation) yielded consistent results: discordant care HR 1.51 (1.30–1.76), orientation assessed HR 1.37 (1.18–1.59), and night shift HR 1.29 (1.22–1.37), with behavioral variables maintaining consistent effect estimates across specifications.

### Sensitivity Analyses

Model performance was robust across sensitivity analyses. Excluding comfort care patients (N = 43,360) yielded M5 AUROC 0.700, and restriction to SOFA 0 (N = 23,668) yielded 0.719. Both exceeded the full-cohort estimate, consistent with the signal being strongest in the lowest-acuity patients.

#### Sedation confounding cascade

Approximately 79.1% of the orientation log-odds ratio at SOFA 0 was attenuated when deep sedation and mechanical ventilation were included as covariates. However, the discordant care composite signal retained significance after full adjustment (OR 1.27, p = 1.29×10 at SOFA 0, estimated excluding the collinear orientation variable). In the most conservative test—SOFA 0, no deep sedation, no mechanical ventilation (N = 11,158)—orientation omission remained independently associated with mortality (adjusted OR 1.52, p = 0.027). Within deeply sedated SOFA 0 patients, those without orientation assessment still had higher mortality (OR 1.35, p < 0.001). Full sensitivity cascade is presented in Supplementary Table S3.

## Discussion

This study demonstrates that behavioral telemetry—computational analysis of clinician documentation patterns—provides statistically significant incremental predictive value for ICU mortality beyond conventional acuity scores and demographics. The clinical model (M5) incorporating behavioral signals and sedation/ventilation status achieved a ΔAUROC of 0.052 over demographics alone, with all comparisons highly significant by DeLong test. The enriched model (M7) incorporating documentation velocity reached 0.723, an improvement of 0.083 over demographics.

### The Confounding Structure: Sedation as Mediator, Not Confounder

A critical finding is the reversal of the orientation assessment coefficient when deep sedation and mechanical ventilation enter the model. In earlier analyses (Papers 1–3),^3–5^ missing orientation was strongly associated with mortality. Here, when sedation is explicitly controlled, orientation assessed becomes positively associated with mortality (OR 1.47). This is not contradictory—it reveals the confounding structure.

Deep sedation and mechanical ventilation are best understood as mediators on the causal pathway—clinical deterioration leads to sedation, which prevents orientation assessment—rather than traditional confounders. As Schisterman and colleagues have demonstrated, adjusting for mediators introduces over-adjustment bias that attenuates the total effect.^15^ The OR of 4.29–5.65 reported in Paper 1 represents the total effect including the sedation-mediated pathway. The OR of 1.27–1.52 reported here after full sedation adjustment represents the direct effect—the component not mediated through sedation.

Three findings support the mediator interpretation. First, approximately 79% of the orientation log-odds ratio at SOFA 0 is explained by the sedation pathway, consistent with mediation rather than confounding. Second, the discordant care composite retains significance (OR 1.27, p < 0.001; estimated excluding the collinear orientation variable) after full adjustment, indicating a behavioral signal independent of sedation status. Third, in the cleanest possible subgroup— SOFA 0, no sedation, no ventilation (N = 11,158)—orientation omission still predicts mortality (OR 1.52, p = 0.027). A true confounder would eliminate the signal entirely; a mediator would attenuate it, which is exactly what we observe. Cox proportional hazards analysis confirmed these findings in a time-to-event framework, with discordant care HR 1.49 (concordance 0.67) and the behavioral variables satisfying the proportional hazards assumption.

Importantly, the discordant care flag captures a qualitatively different signal from orientation omission alone: patients receiving extensive routine physical care WITHOUT cognitive engagement, regardless of sedation status. The combination of high routine care documentation velocity with absent orientation assessment encodes a pattern of clinical attention that may reflect unrecognized cognitive deterioration or missed delirium.^8^

### More Care, Wrong Care

The documentation velocity analysis definitively refutes the neglect hypothesis. If orientation omission simply reflected undertriaged or neglected patients, we would expect lower overall documentation. Instead, discordant patients received 30.5% MORE chart events, with excess documentation in respiratory (+101%), care plan (+94%), and treatment (+22%) categories. The sole deficit was neurological assessments (−15.4%)—precisely the category that includes cognitive evaluation.

This “more care but wrong care” pattern suggests a systematic mismatch: clinicians are actively caring for these patients but directing attention toward physical and respiratory management rather than cognitive assessment. Given that 90.5% of discordant patients had deep sedation and 88.2% were mechanically ventilated, this pattern likely reflects an appropriate focus on ventilator management—but one that may mask evolving delirium or cognitive changes that predict mortality.

### Clinical Implications

The practical implication is that documentation pattern analysis is implementable as a real-time EHR alert. The nine-variable clinical model (M5) uses only data available within the first 24 hours of ICU admission and requires no additional data collection beyond what is already routinely documented. An EHR-integrated system could flag patients exhibiting discordant care patterns for targeted cognitive assessment, potentially creating a treatment window for early intervention.

The compound risk finding is particularly actionable: patients with SOFA 0 + night shift + discordant care had 16.1% mortality—2.6 times the baseline—identifying a specific high-risk subpopulation within an ostensibly low-acuity group. The time-to-first-orientation analysis further supports this: the U-shaped mortality pattern, with lowest mortality among those assessed at 8–12 hours and highest among those never assessed, suggests that timing of cognitive engagement carries independent prognostic information.

### Comparison to Existing Prediction Models

The AUROC values achieved here (0.691–0.723) are modest in absolute terms but reflect the inherent difficulty of mortality prediction in low-acuity patients (SOFA 0–2). Traditional severity scores perform well in high-acuity populations but offer little discrimination when physiologic derangement is minimal. For comparison, the Rothman Index—a nursing assessment-derived composite—achieves AUCs exceeding 0.90 for 24-hour mortality across mixed-acuity populations but incorporates 26 variables including laboratory values not available in our parsimonious 9-variable model.17 Machine learning-based early warning scores such as eCART report AUROCs of 0.74–0.80 for clinical deterioration in ward patients, though these predict ICU transfer rather than mortality in already-admitted ICU patients.18 A systematic review of early warning scores found widespread methodological limitations and questioned whether scores perform as well as reported.19 The behavioral telemetry approach fills this gap by leveraging a complementary information source: clinician behavior patterns rather than physiologic parameters alone. The NRI of 24.3% indicates meaningful reclassification, with approximately one in four patients moving to a more appropriate risk category.

### Limitations

Several limitations warrant discussion. First, this is a single-center retrospective study using MIMIC-IV; external validation across multiple sites is needed. Second, the AUROC values, while statistically significantly improved, remain modest in absolute terms, reflecting the inherent difficulty of mortality prediction in low-acuity patients. Third, 90.5% of discordant patients had deep sedation, creating substantial overlap between the discordant care and sedation phenotypes; however, the discordant care signal survives full sedation adjustment (OR 1.27 at SOFA 0, estimated excluding the collinear orientation variable; see Sensitivity Analyses) and persists in the sedation-free subgroup (OR 1.52), demonstrating that the behavioral component is not merely a proxy for sedation status. Fourth, causation cannot be inferred: documentation patterns may be markers rather than mediators of risk. Fifth, MIMIC-IV uses date-shifted data, precluding analysis of secular trends at calendar resolution. Sixth, CAM-ICU assessment was available for only 2.4% of the cohort, preventing direct delirium adjustment. Seventh, the cohort includes multiple stays per patient; sensitivity analysis restricted to first stays showed consistent results. Full model coefficients for all seven progressive models and the complete variable list are presented in Supplementary Tables S1 and S2.

## Conclusion

Behavioral telemetry signals derived from routine EHR documentation provide statistically significant, clinically meaningful incremental predictive value for ICU mortality in low-acuity patients. The “more care but wrong care” phenomenon—where discordant patients receive more documentation overall but fewer neurological assessments—supports the theoretical framework that documentation patterns encode latent signals of clinical recognition. The sedation confounding structure is consistent with mediation rather than spurious association, and the behavioral signal persists in the cleanest available subgroup. These findings establish the foundation for real-time EHR-based clinical decision support systems that leverage negative telemetry for early risk identification.

## Declarations

### Ethics approval

This study used de-identified data from MIMIC-IV, a publicly available dataset accessed through PhysioNet credentialed access. Institutional review board approval was obtained by the database maintainers (MIT). Individual consent was waived due to retrospective, de-identified data.

### Data availability

MIMIC-IV v3.1 is available through PhysioNet (https://physionet.org/content/mimiciv/). The complete BigQuery SQL query used to derive the study cohort is provided in Supplementary Materials. Analysis code is available from the corresponding author upon request.

**AI disclosure: AI tools (Claude, Anthropic) assisted with statistical code review, literature organization, and manuscript formatting. All scientific content, study design, analysis, interpretation, and conclusions are the sole work of the author.**

**Author contributions (CRediT): Greg Born: Conceptualization, Methodology, Software, Validation, Formal Analysis, Investigation, Data Curation, Writing – Original Draft, Writing – Review & Editing, Visualization, Project Administration.**

## References

1. Johnson AEW, Bulgarelli L, Shen L, et al. MIMIC-IV, a freely accessible electronic health record dataset. Sci Data. 2023;10:1.

2. Vincent JL, Moreno R, Takala J, et al. The SOFA (Sepsis-related Organ Failure Assessment) score to describe organ dysfunction/failure. Intensive Care Med. 1996;22:707–710

3. Born G. Behavioral telemetry in the ICU: missing orientation assessment predicts mortality in patients with low acute physiologic derangement. medRxiv. 2026. [preprint]

4. Born G. Discordant care in the ICU: routine nursing assessment without cognitive engagement predicts mortality. medRxiv. 2026. [preprint]

5. Born G. Screening 17 nursing assessments for mortality association: orientation is the only assessment where omission predicts higher mortality. Int J Nurs Stud. 2026. [under review]

6. Born G. Night shift amplifies the discordant care mortality signal in the ICU. BMJ Qual Saf. 2026. [under review]

7. Newman-Toker DE, Schaffer AC, Yu-Moe CW, et al. Serious misdiagnosis-related harms in malpractice claims. Diagnosis. 2019;6:227–240.

8. Ely EW, Shintani A, Truman B, et al. Delirium as a predictor of mortality in mechanically ventilated patients in the ICU. JAMA. 2004;291:1753–1762.

9. Aiken LH, Clarke SP, Sloane DM, et al. Hospital nurse staffing and patient mortality, nurse burnout, and job dissatisfaction. JAMA. 2002;288:1987–1993.

10. Kalisch BJ, Landstrom GL, Hinshaw AS. Missed nursing care: a concept analysis. J Adv Nurs. 2009;65:1509–1517.

11. Collins GS, Reitsma JB, Altman DG, Moons KGM. Transparent reporting of a multivariable prediction model for individual prognosis or diagnosis (TRIPOD). Ann Intern Med. 2015;162:55–63.

12. DeLong ER, DeLong DM, Clarke-Pearce DL. Comparing the areas under two or more correlated receiver operating characteristic curves. Biometrics. 1988;44:837–845.

13. Pencina MJ, D’Agostino RB, Steyerberg EW. Extensions of net reclassification improvement calculations to measure usefulness of new biomarkers. Stat Med. 2011;30:11–21.

14. Hosmer DW, Lemeshow S. Applied Logistic Regression. 2nd ed. Wiley; 2000.

15. Schisterman EF, Cole SR, Platt RW. Overadjustment bias and unnecessary adjustment in epidemiological studies. Epidemiology. 2009;20(4):488–495.

16. Singh H, Giardina TD, Meyer AND, et al. Types and origins of diagnostic errors in primary care settings. JAMA Intern Med. 2013;173:418–425.

17. Rothman MJ, Rothman SI, Beals J. Development and validation of a continuous measure of patient condition using the electronic medical record. J Biomed Inform. 2013;46:837–848.

18. Churpek MM, Yuen TC, Winslow C, et al. Multicenter comparison of machine learning methods and conventional regression for predicting clinical deterioration on the wards. Crit Care Med. 2016;44:368–374.

19. Gerry S, Bonnici T, Birks J, et al. Early warning scores for detecting deterioration in adult hospital patients: systematic review and critical appraisal of methodology. BMJ. 2020;369:m1501.

20. Ely EW, Margolin R, Francis J, et al. Evaluation of delirium in critically ill patients: validation of the Confusion Assessment Method for the Intensive Care Unit (CAM-ICU). Crit Care Med. 2001;29:1370–1379.

