## Supplementary Files for "Behavioral Telemetry for ICU Mortality Prediction: Documentation Pattern Analysis in 46,002 Low-Acuity MIMIC-IV Patients"

**Supplementary Materials**

Behavioral Telemetry as a Predictive Framework for ICU Mortality

Greg Born, MBA

### Table S1. Complete Variable Dictionary

*All variables used in the study with descriptions, data types, sources, and model membership. N = 46,002 ICU stays from MIMIC-IV v3.1.*

| **Variable Name** | **Description** | **Type / Range** | **Source** | **Used In** |
| --- | --- | --- | --- | --- |
| died | In-hospital mortality | Binary (0/1) | MIMIC-IV discharge disposition | All models (outcome) |
| sofa | SOFA score (first 24h) | Integer (0-2) | MIMIC-IV derived, first 24h max | M1-M7 |
| age | Age at ICU admission | Continuous (years) | MIMIC-IV patients table | M2-M7 |
| male | Sex (male) | Binary (0/1) | MIMIC-IV patients table | M2-M7 |
| charlson | Charlson Comorbidity Index | Integer (0-17) | MIMIC-IV derived from ICD codes | M2-M7 |
| has_orientation | Orientation assessment documented | Binary (0/1) | chartevents, first 24h | M3-M7 |
| discordant_care | Discordant care flag | Binary (0/1) | Derived: routine_care_score >= 6 AND has_orientation = 0 | M3-M7 |
| night_shift | Night shift admission (19:00-07:00) | Binary (0/1) | MIMIC-IV admittime | M4-M7 |
| has_deep_sedation | Deep sedation (any RASS <= -3) | Binary (0/1) | chartevents RASS, first 24h | M5-M7 |
| mechanically_ventilated | Mechanical ventilation in first 24h | Binary (0/1) | procedureevents/chartevents | M5-M7 |
| routine_care_score | Routine care score (0-14) | Integer | Sum of 14 nursing assessment flags | M6-M7 |
| has_rass | RASS assessment documented | Binary (0/1) | chartevents, first 24h | M6-M7 |
| has_gcs | Glasgow Coma Scale documented | Binary (0/1) | chartevents, first 24h | M6-M7 |
| has_turn | Turning/repositioning documented | Binary (0/1) | chartevents, first 24h | M6-M7 |
| has_skin | Skin assessment documented | Binary (0/1) | chartevents, first 24h | M6-M7 |
| has_braden | Braden scale documented | Binary (0/1) | chartevents, first 24h | M6-M7 |
| total_chartevents_24h | Total chart events in first 24h | Integer | chartevents count | M7 |
| unique_items_documented_24h | Unique item types documented | Integer | Distinct itemid count | M7 |
| vital_sign_events | Vital sign documentation events | Integer | chartevents category filter | M7 |
| neuro_events | Neurological documentation events | Integer | chartevents category filter | M7 |
| respiratory_events | Respiratory documentation events | Integer | chartevents category filter | M7 |
| has_cough | Cough/deep breathe documented | Binary (0/1) | chartevents, first 24h | Cohort derivation |
| has_lung_sounds | Lung sounds assessment documented | Binary (0/1) | chartevents, first 24h | Cohort derivation |
| has_edema | Edema assessment documented | Binary (0/1) | chartevents, first 24h | Cohort derivation |
| has_abdominal | Abdominal assessment documented | Binary (0/1) | chartevents, first 24h | Cohort derivation |
| has_temp | Temperature assessment documented | Binary (0/1) | chartevents, first 24h | Cohort derivation |
| has_hob | Head of bed documented | Binary (0/1) | chartevents, first 24h | Cohort derivation |
| has_activity | Activity/mobility documented | Binary (0/1) | chartevents, first 24h | Cohort derivation |
| has_pain | Pain assessment documented | Binary (0/1) | chartevents, first 24h | Cohort derivation |
| hours_to_death | Hours from ICU admit to death | Float | MIMIC-IV derived | Survival analysis |
| days_to_death | Days from ICU admit to death | Float | Derived from hours_to_death | Survival analysis |
| chartevents_per_hour | Chart events per hour | Float | Derived | Descriptive |
| cam_icu_assessed | CAM-ICU assessment performed | Binary (0/1) | chartevents | Descriptive |
| cam_icu_positive | CAM-ICU positive (delirium) | Binary (0/1) | chartevents | Descriptive |
| cam_ms1_acute_onset | CAM feature 1: acute onset | Binary (0/1) | chartevents | Descriptive |
| cam_ms2_inattention | CAM feature 2: inattention | Binary (0/1) | chartevents | Descriptive |
| cam_ms3_disorganized | CAM feature 3: disorganized thinking | Binary (0/1) | chartevents | Descriptive |
| cam_ms4_altered_loc | CAM feature 4: altered LOC | Binary (0/1) | chartevents | Descriptive |
| has_direct_delirium_result | Direct delirium result available | Binary (0/1) | chartevents | Descriptive |
| direct_delirium_positive | Direct delirium result positive | Binary (0/1) | chartevents | Descriptive |
| min_rass_24h | Minimum RASS in first 24h | Float (-5 to +4) | chartevents | Descriptive |
| max_rass_24h | Maximum RASS in first 24h | Float (-5 to +4) | chartevents | Descriptive |
| avg_rass_24h | Mean RASS in first 24h | Float | chartevents | Descriptive |
| rass_count_24h | Number of RASS assessments | Integer | chartevents count | Descriptive |
| hours_to_first_orientation | Hours to first orientation doc | Float | chartevents, time delta | Time-to-orientation analysis |
| events_h0_4 | Chart events hours 0-4 | Integer | chartevents time window | Temporal analysis |
| events_h4_8 | Chart events hours 4-8 | Integer | chartevents time window | Temporal analysis |
| events_h8_12 | Chart events hours 8-12 | Integer | chartevents time window | Temporal analysis |
| events_h12_16 | Chart events hours 12-16 | Integer | chartevents time window | Temporal analysis |
| events_h16_20 | Chart events hours 16-20 | Integer | chartevents time window | Temporal analysis |
| events_h20_24 | Chart events hours 20-24 | Integer | chartevents time window | Temporal analysis |
| discharge_location | Discharge location (detailed) | Categorical | MIMIC-IV admissions | Discharge analysis |
| discharge_category | Discharge category (grouped) | Categorical | Derived from discharge_location | Discharge analysis |
| hospital_los_days | Hospital length of stay (days) | Float | MIMIC-IV derived | Discharge analysis |
| stay_id | Unique ICU stay identifier | Integer | MIMIC-IV icustays | Identifier |
| first_careunit | ICU unit type | Categorical | MIMIC-IV icustays | Cohort description |
| weekend | Weekend admission | Binary (0/1) | Derived from admittime | Cohort description |
| admit_year | Year of admission | Integer (2008-2022) | MIMIC-IV admissions | Cohort description |
| comfort_care | Comfort/palliative care documented | Binary (0/1) | chartevents/procedureevents | Sensitivity analysis |
| omission_score_6 | Omission score >= 6 | Binary (0/1) | Derived from assessment flags | Cohort derivation |
| treatment_events | Treatment documentation events | Integer | chartevents category filter | M7 / descriptive |
| pain_sedation_events | Pain/sedation documentation events | Integer | chartevents category filter | M7 / descriptive |
| care_plan_events | Care plan documentation events | Integer | chartevents category filter | M7 / descriptive |

*Total: 62 unique variables across base cohort (28 columns) and enrichment dataset (40 columns, 3 shared keys). Additional 3 assessment variables (oral care, bowel sounds, peripheral vascular) available in separate file for Paper 3 analyses.*

### Table S2. Full Model Coefficients (M1–M7)

*Odds ratios (95% confidence intervals) from logistic regression on full cohort (N = 46,002). Values shown as OR (95% CI). All p < 0.001 unless otherwise noted. Em dash (—) indicates variable not included in that model. Bold variables are behavioral telemetry signals.*

| **Variable** | **M1** | **M2** | **M3** | **M4** | **M5** | **M6** | **M7** |
| --- | --- | --- | --- | --- | --- | --- | --- |
| Intercept | 0.092 (0.088-0.096) | 0.017 (0.014-0.019) | 0.013 (0.011-0.017) | 0.012 (0.009-0.015) | 0.005 (0.004-0.006) | 0.003 (0.002-0.004) | 0.005 (0.003-0.007) |
| SOFA | 1.180 (1.133-1.228) | 1.111 (1.066-1.157) | 1.080 (1.036-1.125) | 1.092 (1.048-1.138) | 1.045 (1.002-1.090) p=0.039 | 1.054 (1.010-1.099) p=0.015 | 1.108 (1.062-1.156) |
| Age | — | 1.025 (1.023-1.027) | 1.026 (1.024-1.028) | 1.026 (1.024-1.028) | 1.028 (1.026-1.030) | 1.028 (1.026-1.030) | 1.028 (1.026-1.030) |
| Male | — | 0.954 (0.895-1.017) p=0.152 | 0.937 (0.879-0.999) p=0.047 | 0.946 (0.887-1.009) p=0.092 | 0.930 (0.871-0.992) p=0.027 | 0.936 (0.877-0.999) p=0.045 | 0.965 (0.904-1.031) p=0.291 |
| Charlson | — | 1.173 (1.150-1.196) | 1.155 (1.132-1.179) | 1.149 (1.126-1.172) | 1.162 (1.139-1.187) | 1.163 (1.139-1.188) | 1.170 (1.145-1.195) |
| **Orientation assessed** | — | — | 1.051 (0.901-1.225) p=0.530 | 0.986 (0.845-1.150) p=0.855 | 1.467 (1.251-1.720) | 1.098 (0.869-1.386) p=0.434 | 1.088 (0.858-1.379) p=0.485 |
| **Discordant care** | — | — | 1.953 (1.664-2.292) | 1.863 (1.587-2.187) | 1.590 (1.352-1.869) | 1.205 (0.951-1.528) p=0.123 | 1.102 (0.867-1.403) p=0.427 |
| **Night shift** | — | — | — | 1.424 (1.335-1.518) | 1.469 (1.377-1.568) | 1.427 (1.336-1.523) | 1.305 (1.222-1.395) |
| **Deep sedation** | — | — | — | — | 1.503 (1.390-1.625) | 1.558 (1.432-1.695) | 1.548 (1.418-1.689) |
| **Mech. ventilation** | — | — | — | — | 1.916 (1.769-2.074) | 1.898 (1.751-2.058) | 0.872 (0.751-1.012) p=0.071 |
| Routine care score | — | — | — | — | — | 1.164 (1.101-1.231) | 1.123 (1.061-1.188) |
| RASS assessed | — | — | — | — | — | 0.743 (0.664-0.831) | 0.838 (0.747-0.940) p=0.003 |
| GCS assessed | — | — | — | — | — | 0.990 (0.767-1.277) p=0.937 | 0.952 (0.735-1.232) p=0.708 |
| Turning assessed | — | — | — | — | — | 1.385 (1.120-1.713) p=0.003 | 1.261 (1.020-1.560) p=0.033 |
| Skin assessed | — | — | — | — | — | 0.579 (0.426-0.787) | 0.655 (0.481-0.893) p=0.007 |
| Braden assessed | — | — | — | — | — | 1.028 (0.794-1.333) p=0.832 | 1.087 (0.836-1.413) p=0.533 |
| Total chart events | — | — | — | — | — | — | 0.999 (0.999-0.999) |
| Unique items doc. | — | — | — | — | — | — | 0.999 (0.998-1.001) p=0.418 |
| Vital sign events | — | — | — | — | — | — | 1.000 (0.999-1.001) p=0.881 |
| Neuro events | — | — | — | — | — | — | 1.001 (1.001-1.002) |
| Respiratory events | — | — | — | — | — | — | 1.007 (1.007-1.008) |

*Note: Orientation assessed coefficient reverses direction (protective in M3/M4, risk factor in M5) when deep sedation and mechanical ventilation enter the model, reflecting confounding structure discussed in the manuscript. In M7, mechanical ventilation reverses when documentation velocity variables absorb the documentation intensity signal.*

### Table S3. Sensitivity Analysis and Sedation Confounding Cascade

*Progressive restriction of the analytic cohort to isolate behavioral telemetry signal from sedation confounding. All models adjusted for age, sex, and Charlson comorbidity index. *Orientation coded as omission (1 − has_orientation) in row 5, adjusted for age and Charlson only.*

| **Analysis** | **N** | **Deaths** | **Orient. OR (95% CI)** | **Discordant OR (95% CI)** | **Deep Sed. OR (95% CI)** | **Vent. OR** | **AUROC** |
| --- | --- | --- | --- | --- | --- | --- | --- |
| **1. Full cohort (M5)** | 46,002 | 4,295 | 1.47 (1.25-1.72) | 1.59 (1.35-1.87) | 1.50 (1.39-1.63) | 1.92 | 0.691 |
| **2. Excl. comfort care** | 43,360 | 3,304 | 1.41 (1.18-1.69) | 1.63 (1.36-1.95) | 1.65 (1.51-1.80) | 1.73 | 0.700 |
| **3. SOFA 0 only** | 23,668 | 1,981 | 1.06 (0.85-1.33) | 1.34 (1.05-1.71) | 1.38 (1.20-1.60) | 2.62 | 0.719 |
| **4. SOFA 0, discordant adj.** | 23,668 | 1,981 | — | 1.27 (1.12-1.43) | 1.35 (1.20-1.51) | 2.67 | 0.719 |
| **5. SOFA 0, no sed, no vent** | 11,158 | 482 | 1.52* (1.05-2.20) | 1.09 (0.52-2.27) | — | — | 0.654 |

**Key findings from the sedation confounding cascade:**

1. Approximately 79% of the orientation log-odds ratio at SOFA 0 is attenuated when deep sedation and mechanical ventilation are included as covariates, consistent with mediation rather than confounding.

2. The discordant care composite retains independent significance after full adjustment at SOFA 0 (OR 1.27, p = 1.29 × 10⁻⁴), confirming that the behavioral signal survives adjustment for sedation and ventilation status.

3. In the cleanest possible subgroup (SOFA 0, no deep sedation, no mechanical ventilation, N = 11,158), orientation omission remains significantly associated with mortality (OR 1.52, p = 0.027), demonstrating that the signal is not entirely attributable to sedation confounding.

4. Within deeply sedated patients at SOFA 0 (N = 10,543), failure to assess orientation remains associated with nearly 2-fold higher mortality (OR 1.91, p < 0.001), suggesting that sedation alone does not excuse the omission of cognitive engagement assessment.

### Supplementary Figure S1. TRIPOD Participant Flow Diagram

**MIMIC-IV v3.1 Database**

Beth Israel Deaconess Medical Center, 2008–2022

↓

**Inclusion Criteria Applied:**

(1) Age ≥ 18 years

(2) SOFA score 0–2 within first 24 hours

(3) Non-neurological ICU unit

(4) ICU length of stay ≥ 24 hours

↓

**Base Cohort: N = 46,004**

↓ Merge with enrichment data (2 stays unmatched)

**Final Analytic Cohort: N = 46,002**

Deaths: 4,295 (9.34%) | Survivors: 41,707 (90.66%)

Discordant care: 8,891 (19.3%) | Concordant care: 37,111 (80.7%)

↓	↓

**SOFA Distribution:** SOFA 0: 23,668 (51.5%) | SOFA 1: 14,233 (30.9%) | SOFA 2: 8,101 (17.6%)

**ICU Units:** MICU 10,534 | MICU/SICU 8,158 | SICU 7,624 | CVICU 7,164 | TSICU 6,170 | CCU 6,152 | Other 200

**Sensitivity Analysis Subgroups:**

Excluding comfort care: N = 43,360 (Deaths: 3,304, 7.6%)

SOFA 0 only: N = 23,668 (Deaths: 1,981, 8.4%)

SOFA 0, no deep sedation, no mechanical ventilation: N = 11,158 (Deaths: 482, 4.3%)

Within deeply sedated at SOFA 0: N = 10,543

Comfort care patients excluded from sensitivity: N = 2,642 (5.7%)
